# A Pipeline for the Automatic Identification of Randomized Controlled Oncology Trials and Assignment of Tumor Entities Using Natural Language Processing

**DOI:** 10.1101/2024.07.01.24309767

**Authors:** Paul Windisch, Fabio Dennstädt, Carole Koechli, Robert Förster, Christina Schröder, Daniel M. Aebersold, Daniel R. Zwahlen

**Affiliations:** Department of Radiation Oncology, Cantonal Hospital Winterthur, Winterthur, Switzerland; Department of Radiation Oncology, Inselspital, Bern University Hospital, University of Bern, Bern, Switzerland

**Keywords:** Natural language processing, Randomized controlled trial, Evidence-based medicine, Oncology, Metastases, Machine Learning, Transformer, Tumor Entity

## Abstract

**Background:** Most tools trying to automatically extract information from medical publications are domain agnostic and process publications from any field. However, only retrieving trials from dedicated fields could have advantages for further processing of the data.

**Methods:** We trained a transformer model to classify trials into randomized controlled trials (RCTs) vs. non-RCTs and oncology publications vs. non-oncology publications. We assessed the performance and then developed a simple set of rules to extract the tumor entity from the retrieved oncology RCTs.

**Results:** On the unseen test set consisting of 100 publications, the model achieved an F1 score of 0.96 (95% CI: 0.92 - 1.00) with a precision of 1.00 and a recall of 0.92 for predicting whether a publication was an RCT. For predicting whether a publication covered an oncology topic the F1 score was 0.84 (0.77 - 0.91) with a precision of 0.75 and a recall of 0.95. The rule-based system was able to correctly assign every oncology RCT in the test set to a tumor entity.

**Conclusion:** In conclusion, classifying publications depending on whether they were randomized controlled oncology trials or not was feasible and enabled further processing using more specialized tools such as rule-based systems and potentially dedicated machine learning models.

## Introduction

The extraction of PICO (patient, intervention, control, outcome) characteristics from randomized controlled trials (RCTs) using natural language processing (NLP) in automated fashion could be used to improve various meta-research processes, from assessing adherence to reporting standards over using metadata for filtering trials to automating systematic reviews and meta-analyses.^1,2^

Currently available tools, such as Trialstreamer, are mostly domain-agnostic and process RCTs from any field.^3^ However, only retrieving trials from dedicated fields could have advantages for further processing of the data. For example, if a screening model could robustly identify oncology RCTs, a simple rule-based system could be used to identify the tumor entity. If an oncology RCT has the word “prostate” in its title, the chance of it being a publication covering prostate cancer is very high. In order to deploy a rule-based system to find prostate cancer publications in a list of RCTs from any field, the word “prostate” alone would also retrieve RCTs on benign prostatic hyperplasia, prostatitis and others. To increase the specificity one would have to formulate more complex rules with more specific expressions (“prostate cancer”, “prostate adenocarcinoma”, “prostate tumor”) and potentially word ordering (“adenocarcinoma of the prostate”). Formulating bespoke rules for every tumor entity would drastically increase the complexity of the rule-based system which becomes evident when looking at the number of Medical Subject Headings (MeSH) that exist for a given tumor entity.^4^

Another advantage of only retrieving trials from a particular field is that one could pass on the retrieved trials to subsequent domain-specific models that extract additional information, such as the tumor stages that were eligible for inclusion in the trial.^5^ The extracted information should ideally be formalized as a precisely defined question, paired with a list of allowable responses so that it can be used as a common data element (CDE) to ensure consistent data collection in various scenarios.^6^

To test the feasibility of our approach, we developed a pipeline to classify trials into RCTs vs. non-RCTs and oncology publications vs. non-oncology publications and to use a set of rules to subsequently extract the tumor entity from the relevant trials.

## Methods

A random sample of 900 publications from seven major journals (British Medical Journal, JAMA, JAMA Oncology, Journal of Clinical Oncology, Lancet, Lancet Oncology, New England Journal of Medicine) published between 2010 and 2022 were annotated.

Publications that described randomized controlled trials (RCTs) received the label “RCT”. Publications that covered oncological topics received the label “ONCOLGY”. Trials that fulfilled both criteria were assigned both labels. Trials that were neither RCTs nor covered oncology topics were assigned no label. For the purpose of this prototype, publications on benign tumors such as uterine fibroids were considered oncology publications, due to the similarity of terminology. Annotation was based on the title and abstract, which were retrieved as a txt file from PubMed and parsed using regular expressions.

100 randomly sampled trials from the New England Journal of Medicine were used as the unseen test set as the journal publishes both oncology and non-oncology articles. We decided against taking a random sample of all trials as the test set since the model might learn properties of the oncology-focused journals (JAMA Oncology, Lancet Oncology, and Journal of Clinical Oncology) during training. This would improve the performance of the model on the test set but does not generalize to the real-world application with the model not knowing beforehand if a journal is a dedicated oncology journal.

The remaining 800 trials were used to train and validate a multilabel text classification model using a random 85:15 split. The transformer model RoBERTa-base was trained using Adam as the optimizer.^7,8^ The detailed configuration file with all parameters used for training and validation is available from the code repository at https://github.com/windisch-paul/oncology_pipeline.

For testing, a threshold of 0.5 was used to assign predictions to a class. 95% confidence intervals were estimated using normal approximation intervals. Training, validation, and testing were performed in python (version 3.11.5) using, among others, the pandas (version 2.1.0), spacy (version 3.7.4), and spacy-transformers (1.2.5) packages.

For the rule-based entity assignment system, we iteratively developed regular expressions that matched certain strings or substrings in the title of an article (e.g., “prostat”) and assigned publications to a tumor entity based on the presence or absence of these matches. After assigning the entities, publications were grouped into specialties based on the entities (e.g. articles that were assigned “prostate cancer” were grouped into “urogenital tumors”). All development of regular expressions was done on the training/validation set so that the regular expressions were only deployed on the test set once at the end. A complete list of the tumor entities, regular expressions used for matching, and specialties can be found in the code repository.

## Results

Figure 1 presents the distribution of randomized controlled trials (RCTs) and oncology publications in the training/validation and test set. 43.4% of trials in the training/validation set and 74.0% of trials in the test set were RCTs. 27.5% of trials in the training/validation set and 22.0% of trials in the test set were oncology publications.

**Figure 1.**
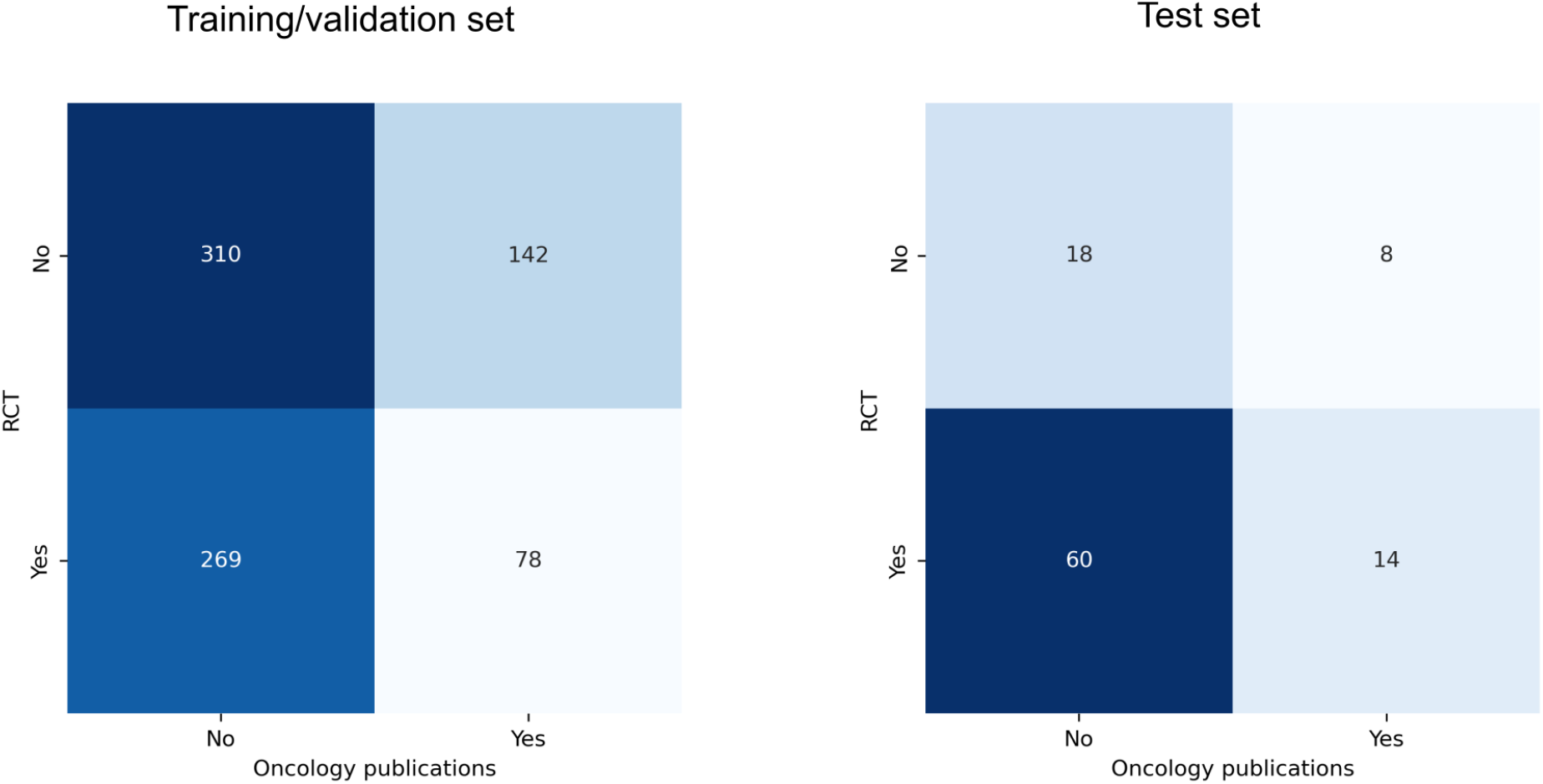
Distribution of trials randomized controlled trials (RCTs) and oncology publications in the training/validation and test set.

The best-performing model during training achieved an F1 score of 0.98 on the validation set when predicting whether a publication was an RCT (precision 1.00, recall 0.96). For predicting oncology publications, the F1 score was also 0.98 (precision 0.96, recall 1.00).

All performance metrics on the test set, including confidence intervals, can be found in Table 1. The confusion matrices are presented in Figure 2.

**Table 1.**
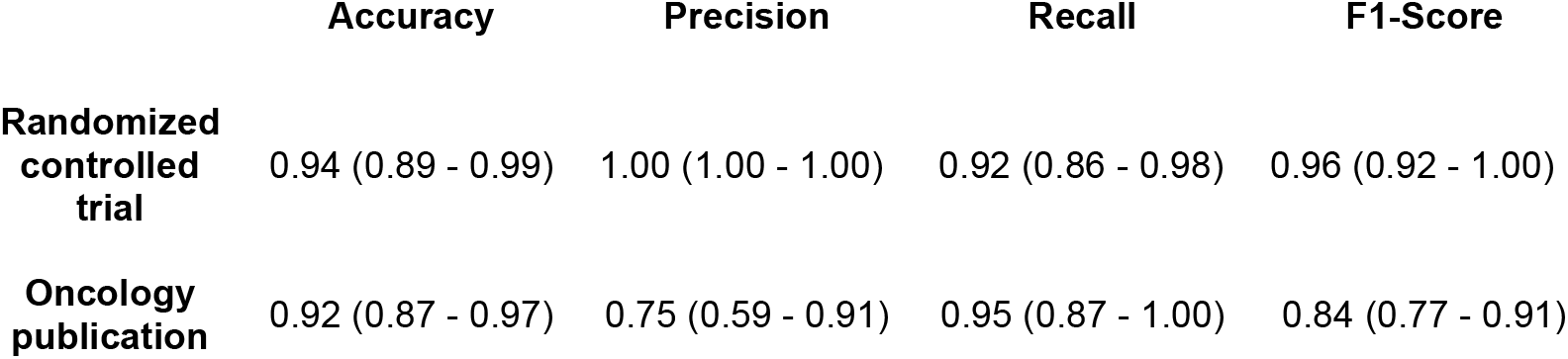
Performance metrics on the test set. Numbers in parentheses indicate the 95% confidence intervals.

|  | Accuracy | Precision | Recall | F1-Score |
| --- | --- | --- | --- | --- |
| **Randomized controlled trial** | 0.94 (0.89 - 0.99) | 1.00 (1.00 - 1.00) | 0.92 (0.86 - 0.98) | 0.96 (0.92 - 1.00) |
| **Oncology publication** | 0.92 (0.87 - 0.97) | 0.75 (0.59 - 0.91) | 0.95 (0.87 - 1.00) | 0.84 (0.77 - 0.91) |

**Figure 2.**
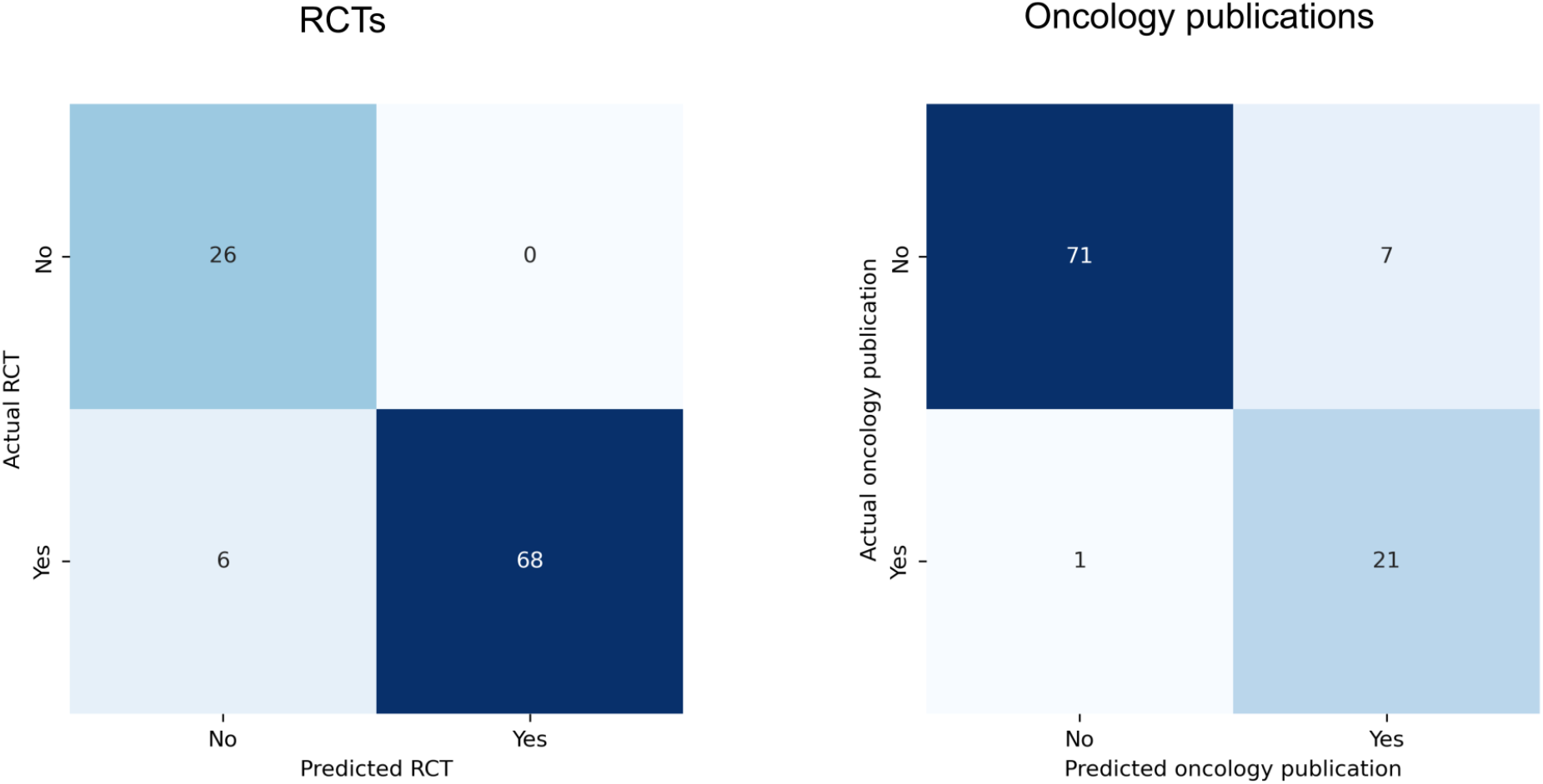
Confusion matrices for the performance of the machine learning model for detecting randomized controlled trials (RCTs - left) and oncology publications (right).

The model achieved accuracies of 0.94 and 0.92 for predicting RCTs and oncology publications, respectively. The precisions were 1.00 and 0.75, the recalls were 0.92 and 0.95, and the F1-scores were 0.96 and 0.84.

The regular expressions were able to assign a tumor entity or metastatic site to 93.6% of oncology RCTs in the training set. The remaining 5 oncology RCTs (6.4%) were either entity agnostic (e.g. investigating remote symptom monitoring for patients with cancer) or enrolled patients with several different entities (e.g. any hematologic malignancy). The same regular expressions were able to assign all 14 oncology RCTs in the test to a tumor entity. The most frequent entities in the training set were non-small cell lung cancer and breast cancer (8.9% respectively). The most frequent entities in the test set were breast and endometrial cancer (21.4 and 14.2% respectively). The distribution of trials by oncology specialty is presented in Table 2. The detailed assignment to the different tumor entities is available from the repository.

**Table 2.** Assignment of trials to oncology specialties in the training/validation set (left) and test set (right). Assignment was performed by iteratively developing regular expressions on the training set and manually reviewing the results.

| Training/validation set |  | Test set |  |
| --- | --- | --- | --- |
| Oncology specialty | n (%) | Oncology specialty | n (%) |
| Gynecological | 16 (23) | Gynecological | 7 (50) |
| Hematological | 14 (20) | Hematological | 3 (21) |
| Gastrointestinal | 13 (18) | Gastrointestinal | 2 (14) |
| Urogenital | 11 (15) | Skin/Soft tissue | 1 (7) |
| Lung/Thoracic | 9 (13) | Urogenital | 1 (7) |
| Skin/Soft tissue | 6 (8) |  |  |
| Head & neck | 2 (3) |  |  |

## Discussion

The proposed transformer models achieved F1 scores of 0.96 when determining if a publication was a randomized controlled trial (RCT) and 0.84 when determining if a publication covered oncology topics. The results regarding RCT classifications are in line with previously published research using various machine learning algorithms such as support vector machines or convolutional neural networks for the same task.^9,10^ The strong performance is especially reassuring considering that the percentage of RCTs was substantially larger in the test set compared to the training/validation set (74.0% and 43.4%, respectively). Incorrect classifications are mainly limited to specific study designs that were relatively rare in the training set, e.g., dose-finding studies with patients randomly assigned to different dosing schedules. For those trials, the terminology around the random assignment and comparisons between different arms might lead the model to conclude that the abstract covers an RCT even though there is no control arm. Exposing the model to examples of these trials, which tend to be more frequent in less prestigious journals, might improve the classification even more in the future.

The oncology classification also achieved good performance that was, however, negatively affected by the precision of 0.75. Here as well, training with additional oncological examples might be helpful as oncology publications only comprise a small percentage of all RCTs. In turn, the model had only seen 220 oncology publications during training and validation, which might be on the lower end in order to get a concept of the oncology-specific terminology.

The iterative development of regular expressions to classify the oncology RCTs into entities was very simple, which supports the hypothesis that retrieving trials from dedicated fields has advantages when processing the data further.

This study has several limitations. First, we only used trials from seven journals for training and testing. While these are probably the journals that publish most practice-changing RCTs in oncology, we cannot assess the model’s ability to generalize to trials from other journals, especially those that use unstructured abstracts. Indeed, for journals where the frequency of RCTl publications is lower, the performance, especially with regard to false positives, needs to be carefully evaluated. In general, the need for a model that predicts whether a publication covers an RCT could be greatly reduced if adherence to guidelines that require identifying RCTs in the title, such as CONSORT, were enforced more rigorously.^1,11,12^ Another limitation is that confidence intervals were estimated using normal approximation intervals while bootstrapping different training sets would likely have resulted in a more accurate estimate. However, training several hundred transformer models seemed excessive, considering the only marginal gain in information. To enable readers to judge the performance of the rule-based entity classification, a filter based on the regular expressions presented herein can be tested on https://www.scantrials.com/.

The strengths of this study include the use of a dedicated unseen test set and the high degree of reproducibility as all code and annotated data are shared in a public repository. As an outlook, training the model with more examples, e.g., of oncology publications as well as of rarer trial designs, could be an option to improve the performance further. However, even in their current form, models like this can be a support tool when screening the results of a literature search.^13,14^

In conclusion, classifying publications depending on whether they were randomized controlled oncology trials or not was feasible. This approach enables further processing using more specialized tools, such as rule-based systems and potentially dedicated machine learning models.

## Data Availability

All data and code used to obtain this study's results have been uploaded to https://github.com/windisch-paul/oncology_pipeline.

